# A mechanistic statistical model of dengue dynamics in an endemic region

**DOI:** 10.64898/2026.09.01.26361961

**Authors:** Nicolás Luna-Martínez, Erika Ximena Cruz-Rodríguez, Edgar Andrés Bernal-Castro

**Author notes:** Corresponding author: (NLM).

## Abstract

**Background:** Dengue is a major public health challenge, and predictive models are crucial for early warning systems. However, many current modeling practices rely exclusively on climatic factors or employ complex algorithms that lack the interpretability needed for informed public health decision-making. To address these shortcomings, we developed and validated a multidimensional, interpretable statistical model to predict monthly dengue incidence.

**Methodology/Principal Findings:** We used a Generalized Linear Mixed Model (GLMM) with a Negative Binomial distribution to analyze 14 years (2010–2023) of spatiotemporal data from 37 municipalities in Huila, Colombia, an endemic region. The model integrates non-linear and lagged effects of climatic, demographic, and socioeconomic factors. The final model underwent rigorous external validation on an independent test set (2021–2023). Our model demonstrated high predictive discrimination (R² = 0.743, Spearman’s ρ = 0.657), accurately capturing the timing of epidemic outbreaks. Key findings include the identification of an optimal thermal window for transmission at 27–28°C, a threshold effect for precipitation above 800 mm, and a saturation dynamic in outbreak autocorrelation.

**Conclusions/Significance:** This mechanistically-informed statistical approach provides a robust and transparent tool for epidemiological surveillance, successfully balancing high predictive performance with the explanatory power needed for effective, data-driven public health interventions.

**Author Summary:** Predicting dengue outbreaks is a complex challenge. To protect communities, public health authorities need tools that are not only accurate but also understandable. Many advanced prediction models are like ’black boxes,’ making it difficult to see why they are forecasting an increased risk, which hinders effective decision-making.

In our study, we built a statistical model that opens up this black box, identifying and explaining the key drivers behind dengue outbreaks. Using 14 years of data from a high-incidence region in Colombia, we integrated information on climate, geography, and social conditions to create a more complete picture of the disease’s behavior. We found clear patterns, such as an “ideal temperature” for dengue transmission and how periods of intense rainfall can trigger outbreaks.

The main advantage of our model is its transparency. It predicts with good accuracy when an outbreak is likely to start, but more importantly, it helps explain why the risk is changing. This allows health authorities to shift from a reactive to a proactive stance. By using specific insights, like an upcoming period of optimal temperature, they can guide targeted interventions, such as eliminating mosquito breeding sites or launching awareness campaigns, exactly where and when they are needed most.

## Introduction

Dengue is one of the world’s most rapidly spreading mosquito-borne viral diseases, representing a critical public health challenge across tropical and subtropical regions [1]. With an estimated 390 million infections occurring annually, its global burden of morbidity and mortality continues to expand [2]. Although the clinical spectrum ranges from mild febrile illness to severe forms, life-threatening manifestations can develop rapidly without timely diagnosis and clinical management, particularly in resource-constrained settings [3]. Recurrent epidemic waves frequently overwhelm healthcare infrastructure, imposing severe socioeconomic disruption on endemic countries [3]. Consequently, accurate and timely predictive early warning systems are essential to anticipate transmission surges, optimize vector control, and guide the rational allocation of healthcare resources [4–6].

Dengue transmission dynamics operate within a complex system governed by non-linear interactions among human populations, the Aedes aegypti vector, and environmental drivers [3,7,8]. In response, an extensive body of predictive modeling literature has emerged. A recent systematic review of 99 dengue outbreak prediction models revealed that all evaluated frameworks relied on climatic variables [9]. Among these, ambient temperature was the most prevalent predictor (95.2%) [10–12], followed by rainfall (81.0%) [12–14] and relative humidity (77.4%) [15,16]. Methodological approaches vary considerably, spanning classical count regressions such as Poisson models (18.3%), autoregressive time series frameworks (26.7%), and a rapidly growing proportion of machine learning algorithms (39.4%) [9].

Despite these developments, current modeling paradigms exhibit substantial shortcomings. First, there is an overreliance on climatic determinants; 70.7% of published models rely exclusively on meteorological covariates, omitting key socio-demographic determinants that modulate transmission risk, with only 5.2% incorporating demographic factors [9]. Second, the increasing adoption of complex machine learning algorithms often enhances statistical pattern recognition at the expense of mechanistic interpretability [17,18]. These ’black-box’ architectures do not readily quantify the individual, non-linear contributions of specific risk factors or elucidate underlying causal mechanisms [17,18]. This opacity represents a major operational barrier in public health, where decision-makers require not only advance notice of outbreak timing, but also transparent biological and environmental thresholds to deploy targeted interventions effectively [17,18]. Finally, methodological rigor remains suboptimal: 20.2% of published models report no validation, and only 5.2% conduct temporal external validation—an essential benchmark to assess generalizability and real-world utility [9,19,20].

To address these gaps, this study aimed to develop and externally validate a multidimensional, interpretable statistical framework for forecasting monthly dengue incidence. We formulated a Generalized Linear Mixed Model (GLMM) with a Negative Binomial distribution designed to explicitly capture non-linear, lagged relationships across climatic, demographic, and socioeconomic domains while maintaining full parametric transparency. We evaluated this model using a 14-year (2010–2023) longitudinal dataset across all 37 municipalities of Huila, Colombia, a Neotropical territory characterized by marked topoclimatic gradients (375 to 1,900 meters above sea level) and endemic-epidemic transmission. Through independent temporal external validation (2021–2023), we demonstrate that our framework achieves high predictive accuracy while preserving the mechanistic interpretability required for actionable public health surveillance and proactive vector control.

## Methods

### Study Design and Setting

We conducted a retrospective, longitudinal ecological study. The study population comprised all 37 municipalities in the department of Huila, Colombia, covering the period from January 1, 2010, to December 31, 2023. The unit of analysis was the municipality-month, forming a spatiotemporal panel dataset with a total of 6,216 observations. A census-based approach was used, including all available observations for each municipality within the study period. The design, linkage, and reporting of this study conform to the Strengthening the Reporting of Observational Studies in Epidemiology (STROBE) guidelines [21] and its extension for Routinely-collected Data (RECORD) [22].

### Ethics Statement

This study was based on publicly available, anonymized, and aggregated secondary data from official sources. Therefore, specific ethics committee approval was not required.

### Data Sources and Variables

The research utilized secondary data from official public sources. Monthly dengue case counts were obtained from Colombia’s National Public Health Surveillance System (SIVIGILA) [23]. Monthly time series of average temperature and total precipitation were compiled from the hydrometeorological database of the Institute of Hydrology, Meteorology, and Environmental Studies (IDEAM) [24]. Annual population projections and the Unsatisfied Basic Needs (UBN) index were sourced from the National Administrative Department of Statistics (DANE) [25]. Population density for each municipality was calculated using annual population projections and the municipality’s area in square kilometers. The response variable was the monthly count of dengue cases per municipality. Predictor variables included climatic, demographic, and socioeconomic factors, as well as their temporal derivatives.

### Data Processing and Imputation

Since climatic data from meteorological stations had missing values for certain months, we implemented an imputation strategy based on geographical proximity, assuming that nearby stations exhibit similar thermal behaviors. The closest station to the one with missing data was identified using the Haversine distance, a metric that calculates the shortest distance over a sphere’s surface between two points defined by latitude and longitude [26]. The missing temperature value was then imputed by adjusting the value from the nearest station by the historical average difference between the two stations, a method that preserves each station’s unique microclimatic characteristics. The formula used was:

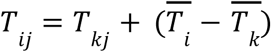

Where *T_ij_* is the imputed temperature for station *i* at time *T_kj_* is the observed temperature at the nearest station *k*, and (*T̄_i_* – *T̄_k_*) is the historical average difference between them. An additional adjustment for altitude was omitted after observing it introduced inconsistencies for mountain stations. For precipitation, a direct imputation from the nearest station was used without a historical average adjustment. This decision was based on precipitation’s high spatial and temporal variability, which does not follow predictable gradients. Using the direct value from the nearest station ensures geographical coherence without introducing the bias that an average adjustment could create for such a fluctuating variable. Finally, to capture the disease’s temporal dynamics, key derived variables were generated: (a) lagged variables for dengue cases and climate data up to three months prior; (b) cumulative variables, such as total precipitation over the previous three months; and (c) a categorical indicator to control for the structural shift during the COVID-19 pandemic (pre/post-March 2020).

### Statistical Analysis

All data analysis was performed using the R programming language (version 4.5.1) with the glmmTMB [27], splines [28], and DHARMa [29] packages. The analysis strategy was divided into four phases: exploratory data analysis (EDA), bivariate analysis, multivariate statistical modeling, and model validation.

Given the nature of the data, overdispersed counts with a hierarchical structure (monthly observations nested within municipalities), we selected a Generalized Linear Mixed Model (GLMM). We fitted a Negative Binomial regression with a logarithmic link function [30–32]. The equation for the final model was:

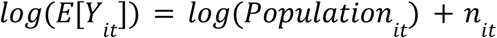

Where *E*[*Y*]*_it_* is the expected number of dengue cases in municipality *i* at time *t*; *log*(*population_it_*) is the offset term to adjust for population size; and *n_it_* is the linear predictor defined as:

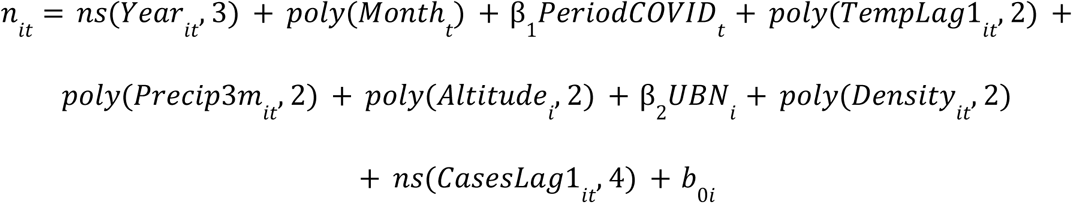

The *offset* term *log*(*population_it_*) allows for modeling incidence rates instead of raw counts, by adjusting the expected number of cases by the population size of each municipality and month. On the other hand, the linear predictive term (*n_it_*) allows for pooling all fixed and random effects. The model’s components include: a natural spline for the non-linear temporal trend (Year); a polynomial transformation for seasonality (Month); a categorical variable for the COVID-19 period; second-degree polynomials for the non-linear effects of lagged temperature (TempLag1) and cumulative precipitation (Precip3m); quadratic polynomials for altitude and population density; a linear term for UBN; and a four-degree-of-freedom natural spline to model the epidemiological autocorrelation (CasesLag1). A random intercept (*b*_0*i*_) for each municipality was included to account for unobserved heterogeneity between territories. This flexible structure, incorporating natural splines [33] and orthogonal polynomials [34], allows for capturing complex non-linear relationships, while the offset and random effects ensure statistical robustness and the correct interpretation of incidence rates in a panel data context.

### Model Selection and Validation

We employed an iterative model-building strategy, progressively adding variables and optimizing functional forms to identify the most robust and parsimonious model. The models were evaluated on a training set (2010–2020) using Akaike’s Information Criterion (AIC) and Bayesian Information Criterion (BIC), and their predictive performance was assessed on an unseen test set (2021–2023) using the coefficient of determination (R²), Root Mean Squared Error (RMSE), Mean Absolute Error (MAE), and Spearman’s rank correlation (⍴).

The final model underwent rigorous diagnostic checks. Goodness-of-fit and model assumptions were evaluated using simulated scaled residuals from the DHARMa package [29]. This analysis confirmed that the Negative Binomial distribution was appropriate for handling overdispersion (dispersion test, p = 0.98), though minor deviations from a uniform distribution were detected (Kolmogorov-Smirnov test, p < 0.001). Additionally, multicollinearity was ruled out, as all predictors had a Variance Inflation Factor (VIF) below 5. To assess predictive performance, we conducted a temporal external validation.

## Results

### Descriptive Analysis and Dengue Incidence Patterns

The distribution of monthly dengue cases per municipality during the study period (2010–2023) revealed a marked positive skew and high overdispersion (Fig 1A). The vast majority of observations were concentrated at or near zero, with a long right-hand tail. The median case count was zero, indicating that no dengue cases were reported in more than half of the municipality-month observations. In contrast, the mean case count was higher (6.381), driven by sporadic but high-intensity epidemic outbreaks, which reached a maximum of 937 cases in a single month for one municipality. This pattern is characteristic of count data for rare epidemiological events.

**Fig 1.**
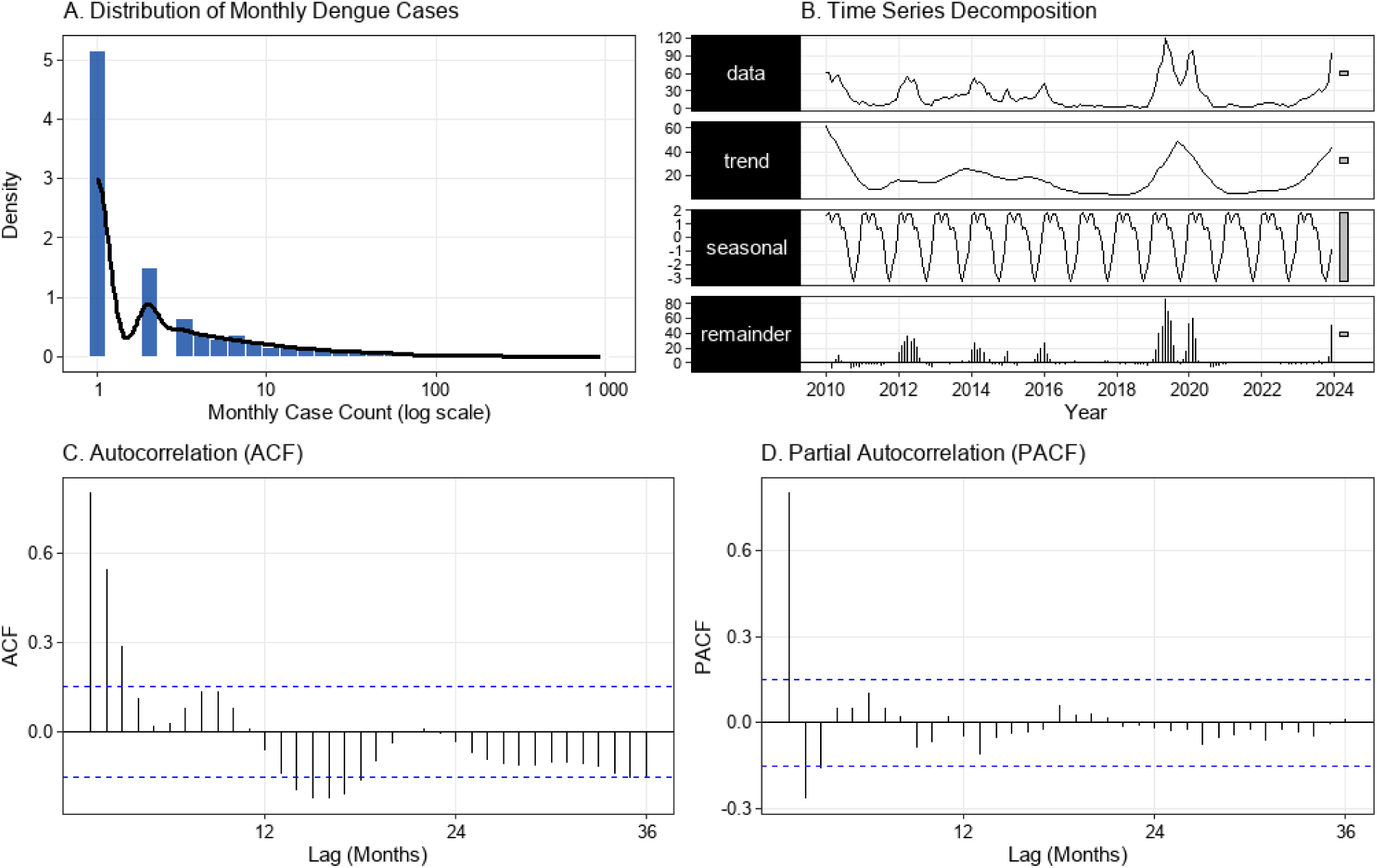
Descriptive and time series patterns of dengue incidence in Huila, Colombia (2010–2023). **(A)** Histogram and density curve of the distribution of monthly dengue cases on a log (+1) scale, showing a marked positive skew with a high concentration of zero-count observations, confirming data overdispersion. **(B)** Seasonal-Trend-Loess (STL) decomposition of the aggregated monthly dengue incidence rate for the department, revealing inter-annual cycles, a strong seasonal pattern, and residual outbreak peaks. **(C)** Autocorrelation Function (ACF) of the residuals, showing a significant correlation at lag 1. **(D)** Partial Autocorrelation Function (PACF) of the residuals, confirming a strong autoregressive pattern of order 1.

The distributions of the main predictor variables also showed distinct patterns. The mean monthly temperature (mean 22.6°C, range 17.6–32.3°C) exhibited a trimodal distribution, suggesting the existence of at least three different thermal regimes within the department. Total monthly precipitation (mean 133.6 mm, max 919 mm) showed a strong positive skew, reflecting the occurrence of intense rainfall events. The distribution of altitude (range 375–1900 masl) was not uniform but clustered, reflecting the different geographical zones of Huila. Finally, the Unsatisfied Basic Needs (UBN) Index (range 7.6–26.4%) revealed a bimodal pattern, indicating that municipalities are grouped into two distinct socioeconomic categories (see S1 Fig for visual distributions).

Analysis of the aggregated time series for the department of Huila revealed well-defined temporal patterns (Fig 1B). The long-term trend showed the cyclical, inter-annual nature of the disease, with periods of high epidemic activity in 2010, 2013–2014, 2016, and a major outbreak in 2019–2020. The trend also showed a resurgence from 2018, followed by a marked decline during the pandemic period (2021–2022) and a new reactivation in 2023. The seasonal component showed a robust, repetitive annual pattern: transmission tends to be lower at the beginning of the year, increases to a peak between May and October, and declines toward December. The residual component highlighted the months with abnormally explosive outbreaks, confirming the intensity of the 2014 and 2019 peaks.

Furthermore, an autocorrelation analysis of the incidence series residuals, after adjusting for trend and seasonality, confirmed a strong temporal dependence (Fig 1C and 1D). The Autocorrelation Function (ACF) showed a very high and statistically significant peak at lag 1 (rho ≈ 0.8), indicating that the residual of one month is strongly correlated with that of the previous month. The Partial Autocorrelation Function (PACF) showed a single dominant and significant peak at lag 1, with correlations for subsequent lags dropping abruptly within the confidence bands. This combined pattern, a decaying ACF and a PACF that cuts off after lag 1, is the classic signature of an autoregressive process of order 1 (AR(1)). This confirms that the number of cases in one month is a strong predictor of the number of cases in the following month, even after controlling for trend and seasonality.

### Selection of the Final Predictive Model

To identify the most robust and parsimonious predictive model, we employed an iterative building strategy, starting with a base model and progressively adding new predictors and more complex functional forms. Table 1 presents the structure of the six models developed in this process, showing the evolution from the simplest to the most complex model.

**Table 1.**
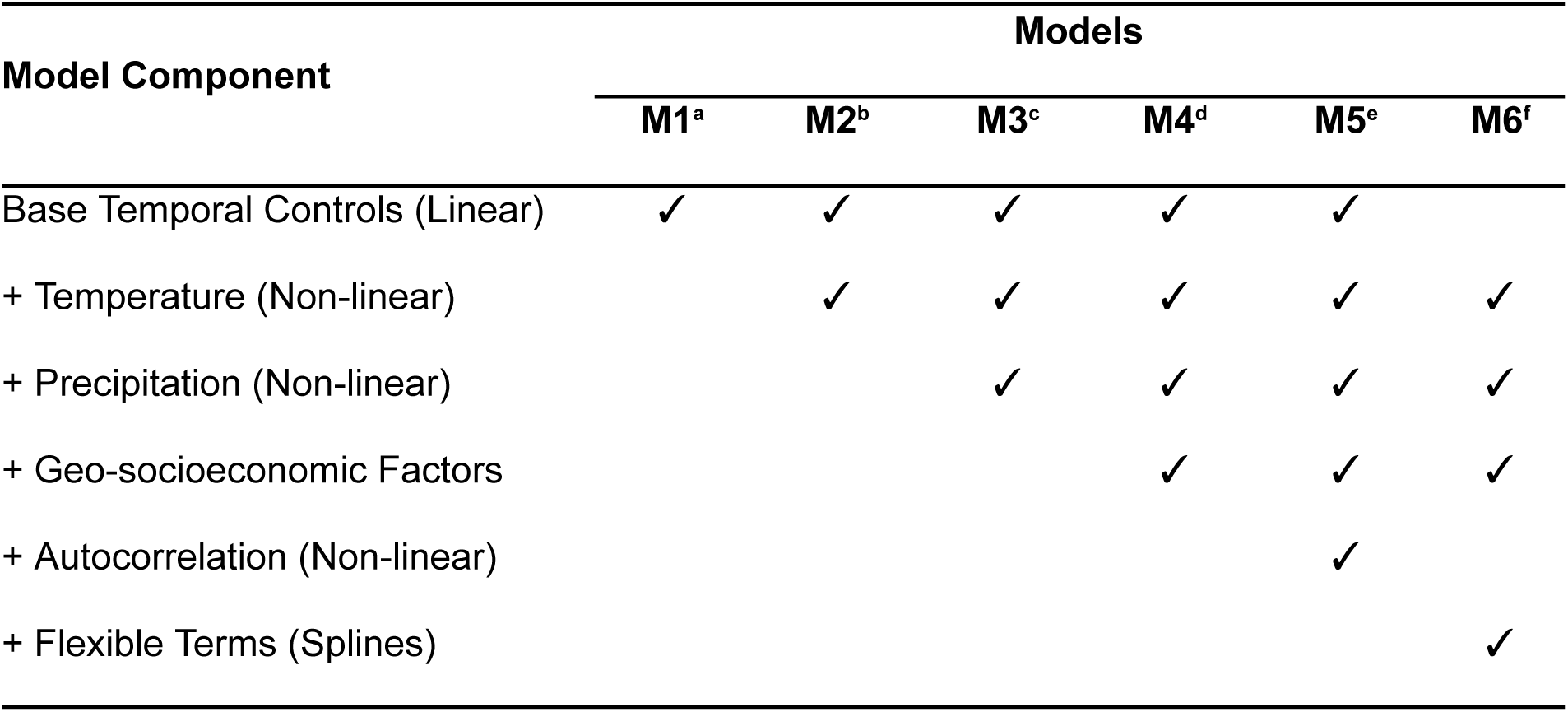
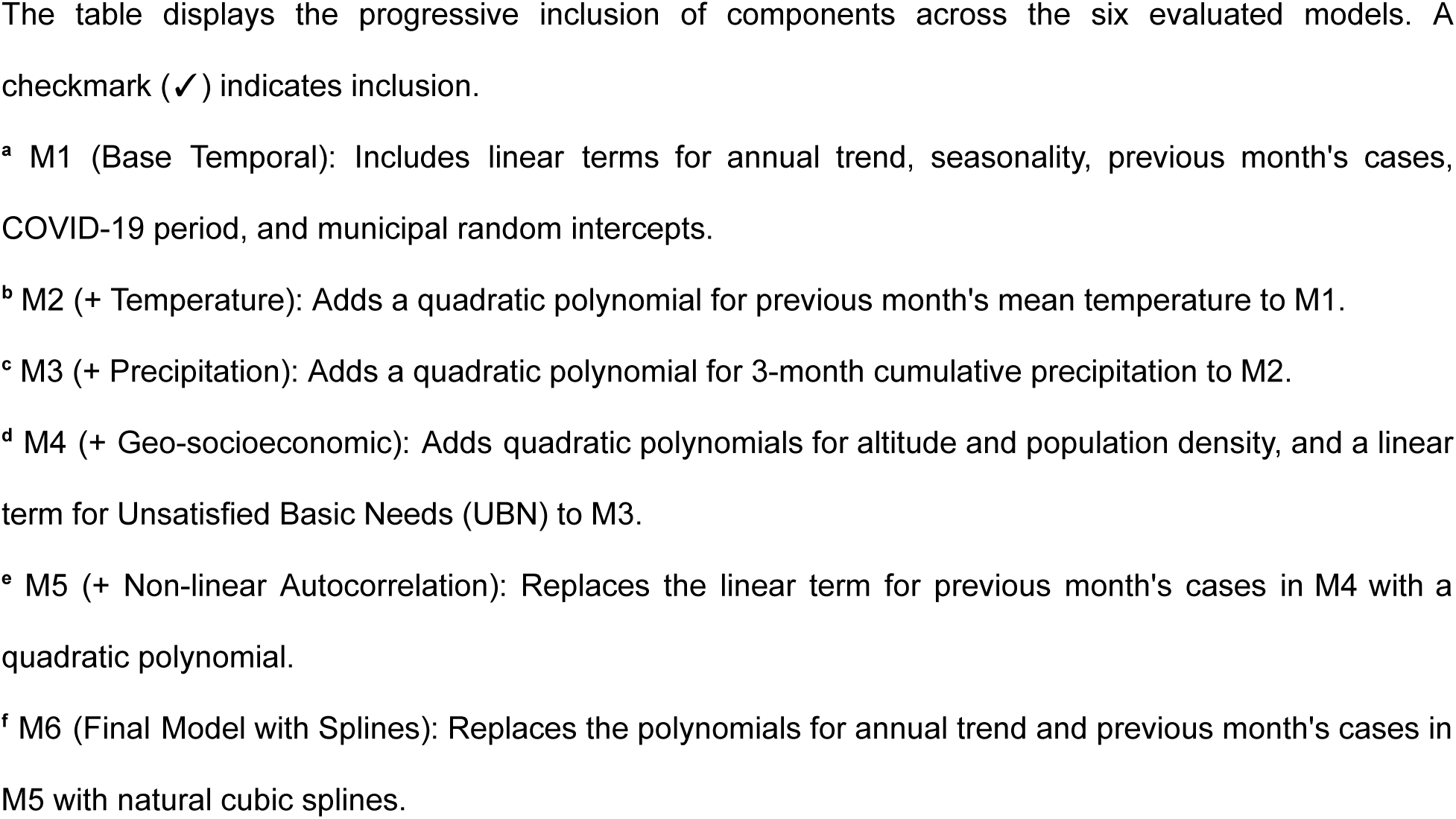
Structure and components of the iteratively developed models.

The performance of each model is detailed in Table 2. The analysis reveals a clear progression as components were added. Including non-linear terms for climatic variables (M2, M3) improved the model’s ability to capture the direction of outbreaks, as seen by the increase in Spearman’s Correlation. However, this came at the cost of introducing prediction instability and increasing error (RMSE and MAE). A qualitative leap occurred when geo-socioeconomic variables were integrated (M4), which drastically reduced the predictive error by almost 90% compared to the purely climatic models.

**Table 2.**
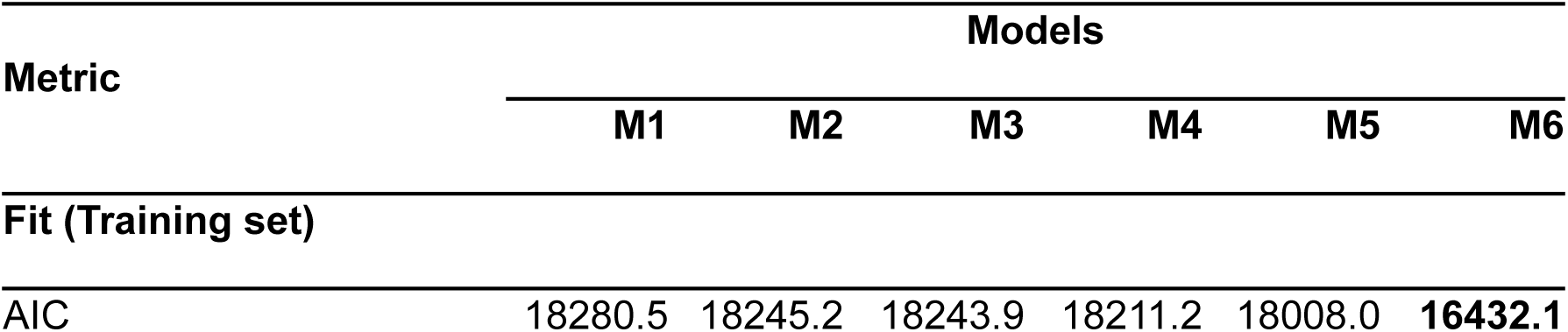
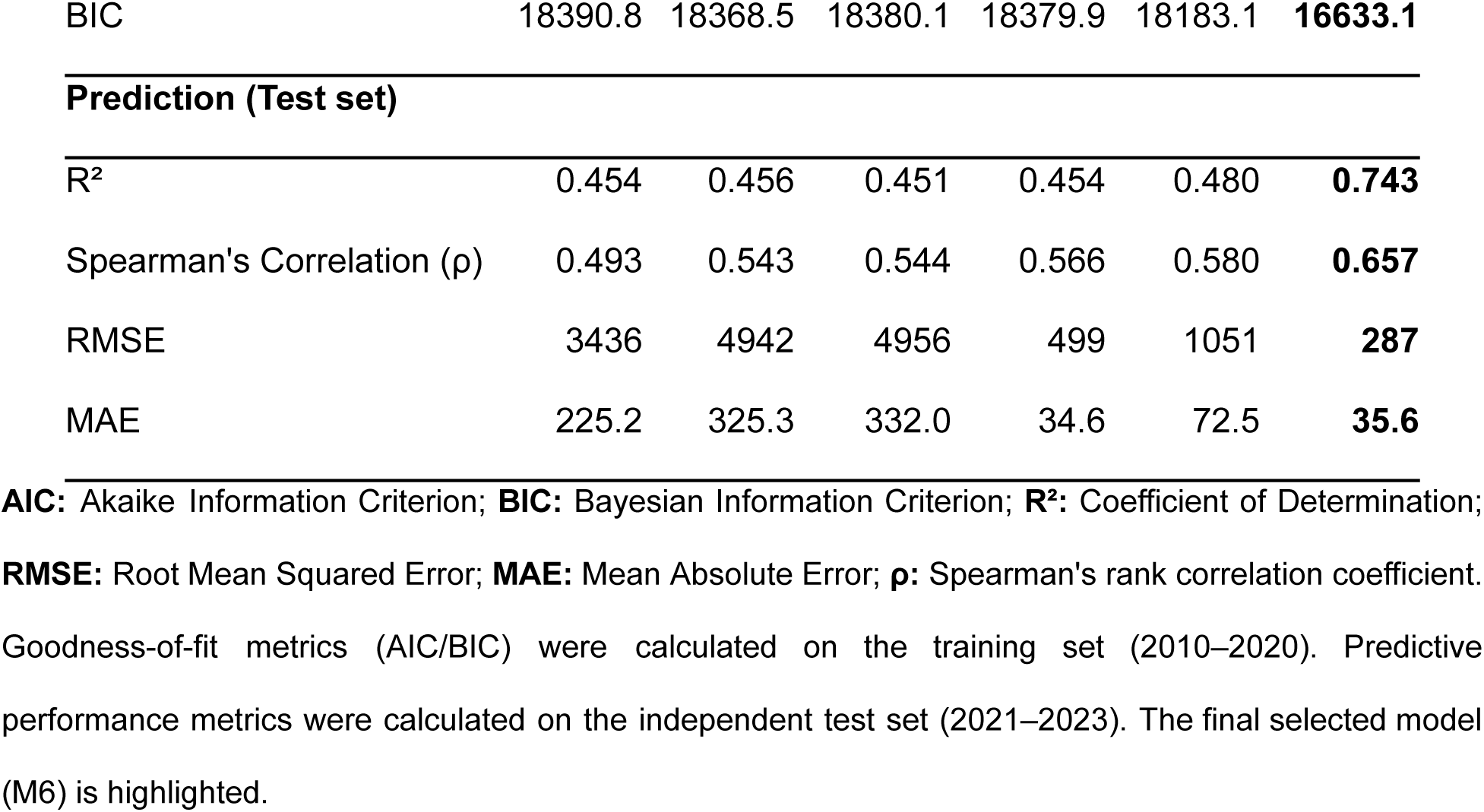
Performance comparison of the iteratively developed models.

Subsequently, while adding a quadratic term for autocorrelation (M5) slightly improved the R², it also doubled the error by generating extreme and unrealistic predictions. This dilemma was resolved with the final model (M6), which replaced the rigid polynomials with more flexible and stable natural splines to model the annual trend and autocorrelation. As shown in Table 2, the M6 model not only had the best statistical fit (lowest AIC/BIC) but also demonstrated the highest and most balanced predictive performance across all metrics. Therefore, it was selected as the final model for the analysis.

### Factors Associated with Dengue Incidence

The final model quantified the associations between multiple predictors and monthly dengue incidence (Table 3). All included predictors, with the exception of the Unsatisfied Basic Needs (UBN) index, showed a statistically significant association with dengue incidence. The model’s key findings are the complex, non-linear relationships, which are visualized as partial effect plots in Fig 2.

**Fig 2.**
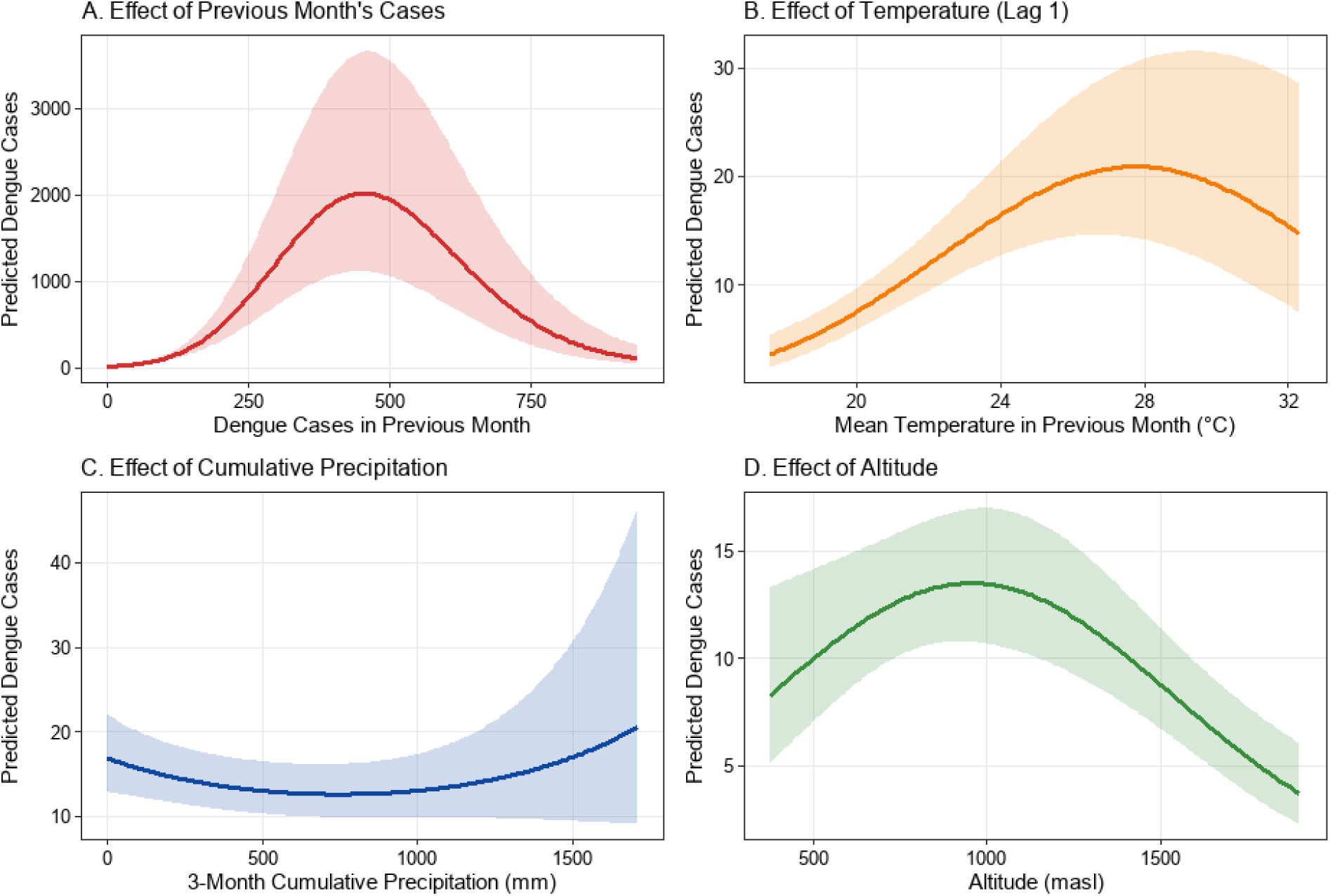
Partial effects of the main predictors from the final model. Each panel shows the marginal effect of a single predictor on the expected monthly dengue case count, holding all other covariates constant at their mean or reference levels. Shaded bands represent 95% confidence intervals. **(A)** Non-linear effect of previous month’s dengue cases (epidemiological autocorrelation). **(B)** Non-linear effect of previous month’s mean temperature, highlighting the optimal thermal transmission window. **(C)** Non-linear effect of 3-month cumulative precipitation, demonstrating the threshold effect. **(D)** Non-linear effect of altitude, showing peak risk at intermediate elevations.

The model captured significant temporal patterns, including a cyclical inter-annual trend (Table 3, p < 0.001) and a substantial reduction in dengue incidence during the COVID-19 pandemic. Controlling for all other factors, the dengue incidence rate was approximately 69.7% lower during the pandemic period compared to the pre-pandemic baseline (Table 3, IRR = 0.303, 95% CI: 0.246 – 0.374).

Climatic factors exhibited significant non-linear relationships with transmission. The previous month’s mean temperature was associated with incidence in an inverted U-shape, revealing an optimal thermal window for transmission (Fig 2B). Similarly, cumulative precipitation over the previous three months exhibited a threshold effect, where risk increased sharply only after exceeding 800 mm of accumulated rainfall (Fig 2C).

Among geo-socioeconomic factors, altitude showed a complex, inverted U-shaped relationship, with peak incidence predicted at mid-range elevations rather than at the lowest altitudes (Fig 2D). In contrast to expectations, population density was negatively associated with dengue incidence (Table 3, p < 0.001). Although the UBN index showed a positive direction of association, it was not statistically significant in the final multivariable model (Table 3, IRR = 1.082, 95% CI: 0.946 – 1.239, p = 0.251). The most influential predictor was epidemiological autocorrelation; previous month’s cases exhibited a strong, inverted U-shaped non-linear association with current incidence, indicating a saturation dynamic in transmission following large outbreaks (Fig 2A).

### Predictive Performance of the Final Model

The final model’s performance was evaluated on an independent test set spanning from January 2021 to December 2023. The model demonstrated strong discriminative ability, explaining 74.3% of the variance in monthly case counts (R² = 0.743) and showing a strong rank correlation between predicted and observed values (Spearman’s ρ = 0.657) (Table 2).

**Table 3.** Results of the final Generalized Linear Mixed Model (M6).

| Predictor | Type | IRR (95% CI) | p-value |
| --- | --- | --- | --- |
| **Temporal Effects** |  |  |  |
| Temporal Trend (Year) | Non-linear (spline) | - | < 0.001 |
| Seasonality (Month) | Non-linear (poly) | - | < 0.001 |
| COVID-19 Period (Ref: Pre-pandemic) | Categorical | 0.303 (0.246, 0.374) | < 0.001 |
| **Climatic Effects** |  |  |  |
| Temperature (Lag 1) | Non-linear (poly) | - | < 0.001 |
| Precipitation (3-month cumulative) | Non-linear (poly) | - | 0.030 |
| **Geo-socioeconomic Effects** |  |  |  |
| Altitude | Non-linear (poly) | - | < 0.001 |
| Population Density | Non-linear (poly) | - | < 0.001 |
| UBN | Linear | 1.082 (0.946, 1.239) | 0.251 |
| **Autocorrelation Effect** |  |  |  |
| Dengue Cases (Lag 1) | Non-linear (spline) | - | < 0.001 |

A visual comparison of the observed and predicted time series confirms the model’s capacity to forecast dengue transmission timing, accurately capturing the onset and trajectory of the 2023 epidemic outbreak (Fig 3).

**Fig 3.**
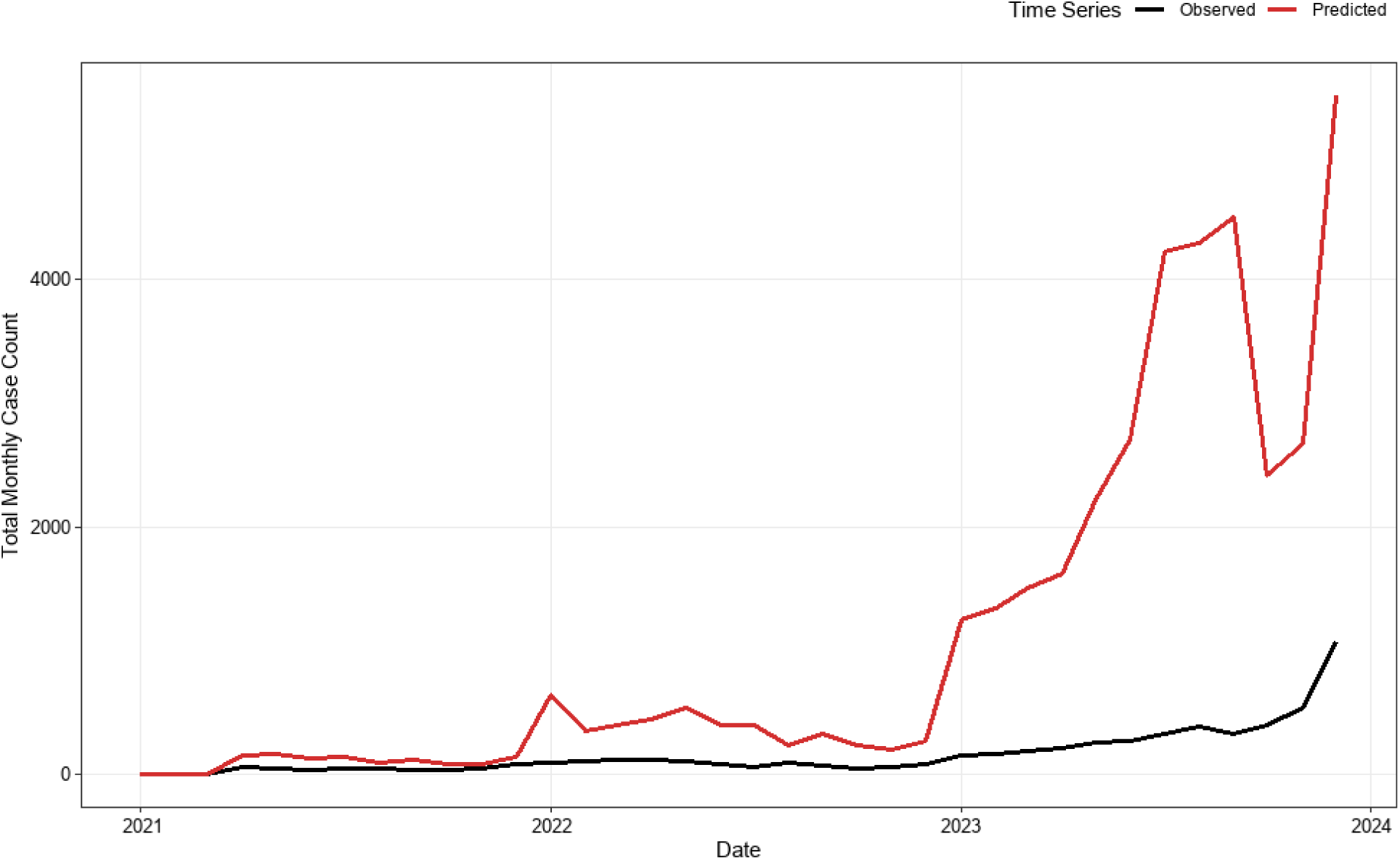
Observed vs. predicted total monthly dengue cases in Huila (2021–2023). The black line represents actual reported cases from SIVIGILA, while the red line represents out-of-sample predictions generated by the M6 model. The model accurately forecasts the timing and inflection of the 2023 outbreak while overestimating its absolute peak magnitude.

## Discussion

This study aimed to develop and validate a multidimensional, interpretable predictive model for dengue incidence. Our results confirm that a Generalized Linear Mixed Model (GLMM) can not only forecast the spatiotemporal dynamics of dengue in an endemic region with high predictive accuracy, but also decipher the complex non-linear relationships that govern disease transmission. Our final, externally validated model demonstrated strong discriminative ability, explaining approximately 74% of the variance in monthly dengue incidence and accurately capturing the timing of epidemic outbreaks. A key contribution of our model is the quantification of non-linear relationships with climatic factors. We identified an optimal thermal window for transmission between 27–28°C, above which transmission risk declines. This finding is consistent with the literature on the thermal biology of the *Aedes aegypti* vector, whose survival, biting rate, and extrinsic incubation period are constrained at extreme temperatures [35–37]. Similarly, the model revealed a threshold effect for cumulative precipitation, where outbreak risk increases sharply only after exceeding 800 mm across the preceding three months. This indicates that it is not isolated rainfall events, but rather the sustained accumulation of water, leading to the stable proliferation of larval breeding habitats, that acts as the primary hydrological trigger for transmission surges [12,38,39]. The most influential predictor was epidemiological autocorrelation; the inverted U-shaped association of previous month’s cases captures the momentum of an outbreak followed by saturation dynamics, likely driven by the localized depletion of susceptible individuals [40] or the intensification of reactive public health vector control interventions after major transmission peaks [41,42], a non-linear biological dynamic that purely linear time series models fail to capture [43].

The analysis of geo-socioeconomic factors yielded nuanced and highly relevant insights. Altitude exhibited a non-linear limiting effect, with peak transmission predicted between 900 and 1,000 meters above sea level, aligning with the established ecological niche of *A. aegypti* in Andean topographies [44]. Counterintuitively, population density was negatively associated with dengue incidence (p < 0.001). While higher host density is theoretically assumed to facilitate transmission, in this subnational context population density likely serves as a proxy for unmeasured urban determinants, such as more continuous piped water access (reducing the necessity for domestic water storage), better infrastructure, or prioritized municipal vector control campaigns in major urban centers [45]. Critically, the Unsatisfied Basic Needs (UBN) index was not statistically significant in the final multivariable model (p = 0.251), suggesting that the baseline socioeconomic effect is largely mediated or confounded by dominant environmental variables such as elevation and microclimate.

In the contemporary landscape of infectious disease forecasting, an ongoing debate exists between mechanistic models, which prioritize causal interpretability, and machine learning algorithms, which optimize predictive accuracy [9,17,18]. Our framework positions itself as an integrative statistical approach, utilizing a regression architecture to explicitly parameterize and test biological transmission hypotheses, thereby aligning with the philosophy of mechanistic models essential for public health decision-making [46,47]. Unlike ’black-box’ machine learning architectures that often lack operational transparency [48], our GLMM allows epidemiologists to not only predict outbreaks but also quantify the functional form and effect size of each covariate. This interpretability is vital for public health practice, enabling authorities to design targeted interventions tailored to the specific environmental and epidemiological drivers dominant at a given time [49–51].

Furthermore, this study directly addresses two critical shortcomings identified in recent systematic reviews. First, it overcomes the unilateral reliance on climatic determinants that characterizes over 70% of existing dengue models [9] by successfully integrating topoclimatic, demographic, and socioeconomic indicators into a unified panel framework. Second, it responds to the imperative for higher methodological rigor; while only 5.2% of published dengue models conduct external validation [9], our framework underwent independent temporal external validation across a three-year period (2021–2023) not involved in model training, providing a transparent assessment of its real-world generalizability.

The evaluation of predictive performance demonstrated strong discriminative capacity (R² = 0.743, Spearman’s ρ = 0.657), confirming the model’s robustness in differentiating between baseline and epidemic periods. Although out-of-sample predictions accurately forecast outbreak timing and acceleration, the model exhibited a tendency to overestimate absolute peak magnitudes during explosive transmission periods (Fig 3). From an epidemiological and early warning standpoint, this characteristic aligns with the Precautionary Principle in public health [52]. A predictive framework that errs on the side of heightened risk during critical transmission windows is operationally safer, as the societal and health system costs of mobilizing preventive resources are markedly lower than the catastrophic burden of an unanticipated epidemic outbreak resulting in delayed response [53,54].

This study possesses notable strengths, including a 14-year longitudinal panel dataset, the integration of multi-source surveillance and climate data, and rigorous out-of-sample temporal validation. Nevertheless, several limitations must be acknowledged. First, as an ecological study, observations represent municipal-level aggregations, and associations cannot be interpreted at the individual level (ecological fallacy). Second, passive surveillance data from SIVIGILA are subject to inherent reporting delays and underreporting of mild or asymptomatic infections. Third, meteorological station data imputation, while geostatistically robust, cannot fully substitute for high-resolution microclimatic sensors. Finally, our models did not account for unmeasured variables such as viral serotype transitions, fine-scale human mobility flows, or localized insecticide spraying coverage.

In conclusion, this work delivers an interpretable, validated predictive model that serves as a robust decision-support tool for dengue surveillance in endemic territories. These findings provide actionable insights for epidemiological preparedness in the department of Huila while establishing a generalizable methodological framework adaptable to other Neotropical regions facing similar arboviral threats. Future research should focus on scaling this framework to national surveillance networks, incorporating real-time climate telemetry, and evaluating hybrid pipelines that integrate statistical transparency with automated machine learning architectures.

## Supporting Information Captions

**S1 Fig. Distribution of the main predictor variables in Huila, Colombia (2010–2023).**

Histograms and density curves for the four main continuous predictor variables used in the statistical models: **(A)** Mean monthly temperature (°C), showing a trimodal thermal distribution; **(B)** Total monthly precipitation (mm), exhibiting positive skewness with extreme rainfall events; **(C)** Municipal altitude distribution (meters above sea level); and **(D)** Unsatisfied Basic Needs (UBN) Index (%), displaying a bimodal socioeconomic pattern across municipalities.

## Data Availability

All data underlying the findings of this study were obtained from publicly available official surveillance and meteorological databases in Colombia. Monthly dengue case counts were sourced from Colombia's National Public Health Surveillance System (SIVIGILA; https://portalsivigila.ins.gov.co/). Climate time series (mean temperature and total precipitation) were compiled from the Institute of Hydrology, Meteorology, and Environmental Studies (IDEAM; http://dhime.ideam.gov.co/). Municipal demographic and socioeconomic indicators (population projections and Unsatisfied Basic Needs index) were retrieved from the National Administrative Department of Statistics (DANE; https://www.dane.gov.co/).

## Notes

### Competing Interest Statement

The authors have declared no competing interest.

### Author Declarations

The study used ONLY openly available, anonymized, and aggregated human and environmental data that were originally located at: 1. Dengue surveillance case counts: National Public Health Surveillance System (SIVIGILA), Instituto Nacional de Salud, Colombia (https://portalsivigila.ins.gov.co/). 2. Demographic projections and Unsatisfied Basic Needs (UBN) index: Departamento Administrativo Nacional de Estadistica (DANE), Colombia (https://www.dane.gov.co/). 3. Hydrometeorological and climate time series: Instituto de Hidrologia, Meteorologia y Estudios Ambientales (IDEAM), Colombia (http://dhime.ideam.gov.co/atencionciudadano/).

