## Supplementary material for "A mechanistic statistical model of dengue dynamics in an endemic region": S1 Fig

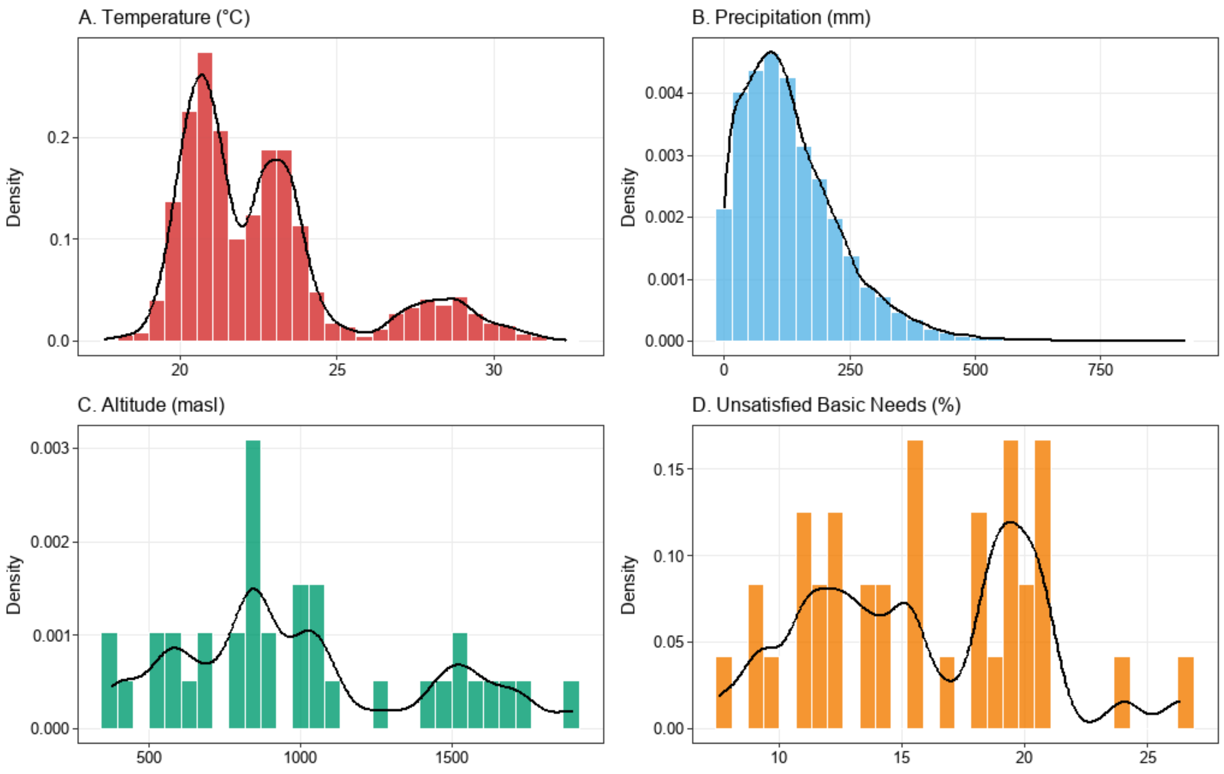


**S1 Fig. Distribution of the main predictor variables in Huila, Colombia (2010–2023).**

Histograms and density curves for the four main continuous predictor variables used in the statistical models: **(A)** Mean monthly temperature (°C), showing a trimodal thermal distribution; **(B)** Total monthly precipitation (mm), exhibiting positive skewness with extreme rainfall events; **(C)** Municipal altitude distribution (meters above sea level); and **(D)** Unsatisfied Basic Needs (UBN) Index (%), displaying a bimodal socioeconomic pattern across municipalities.
